# Seroincidence rate of typhoidal *Salmonella* in children in Kenya

**DOI:** 10.1101/2025.06.24.25330223

**Authors:** Aslam Khan, Polina Kamenskaya, Izabela Rezende, Francis Mutuku, Bryson Ndenga, Zainab Jembe, Priscilla Maina, Philip Chebii, Charles Ronga, Victoria Okuta, Denise O. Garrett, Donal Bisanzio, Kristen Aiemjoy, Jason R. Andrews, A. Desiree LaBeaud, Richelle Charles

## Abstract

Enteric fever, caused by *Salmonella enterica* serovars Typhi and Paratyphi, causes significant morbidity and mortality globally. Surveillance is limited by the lack of reliable diagnostic assays, leading to major gaps in understanding the population-level burden in low- and middle-income countries. We applied a novel serologic tool measuring anti-IgG responses to Hemolysin E (HlyE) to assess infection rates in Kenyan children from four communities: two in western Kenya (Kisumu and Chulaimbo) and two in coastal Kenya (Ukunda and Msambweni). We found a substantially higher enteric fever seroincidence rate in coastal Kenya (37/100 person-years) than in western Kenya (3.6/100 person-years). We found a higher seroincidence rate in households with non-piped water, lower income, and neighborhoods with higher population density. These findings contribute to Kenya’s limited on enteric fever surveillance data, especially in the coastal regions. Such information underscores the need for public health interventions such as typhoid conjugate vaccine introduction in Kenya.

## Introduction

Enteric fever is a significant health problem globally, with the potential to cause a spectrum of symptoms, including severe febrile illness and intestinal perforation [1]. *Salmonella enterica* serovars Typhi and Paratyphi A. B, and C are responsible for the collective term enteric fever, with *S*. Typhi being the most prevalent, followed by *S*. Paratyphi A. Whereas *S*. Typhi is found throughout the world, *S*. Paratyphi A is most prevalent in South and Southeast Asia and not commonly found on the African continent. The majority of illness occurs in low and middle-income countries (LMICs) that lack access to safe drinking water and improved sanitation, with children and adolescents bearing the highest burden of disease [2–4]. Accurate diagnosis is challenging and often requires culture-based methods [5]. The current reference standard is blood culture, which may be unavailable in many clinical settings where typhoid is endemic and has an estimated sensitivity ranging from 51%-65% that varies by age, duration of symptoms, antibiotic use prior to testing, and volume of blood collected [5,6]. Alternative molecular testing using peripheral blood has low diagnostic sensitivity [7]. Point-of-care serologic-based diagnostic tests, like the Widal test, are also limited by low sensitivity and specificity [5]. In addition to variable sensitivity and specificity, available testing for enteric fever may be cost-prohibitive, resulting in underreporting of the true incidence of disease. Furthermore, with public health allocations of resources focused on other infectious diseases, accurate surveillance remains an epidemiological challenge. It is important to capture the burden of these infections to better prioritize preventive measures, including vaccine implementation [3,4].

While geostatistical models estimate a high burden of enteric fever in sub-Saharan Africa, relatively few studies have provided direct disease burden estimates in the region [2,8]. The available data suggest a very high burden of typhoid in Kenya; however, most studies focused primarily on the densely populated capital city of Nairobi and its surrounding areas, with limited data available from the western or coastal regions [2,9]. Although the Kenyan Ministry of Health offers typhoid vaccination for high-risk groups, it has not implemented a routine immunization series for all individuals [9]. To further understand the burden of typhoid in western and coastal Kenya, we implemented a novel serosurveillance tool to estimate the population-level enteric fever seroincidence rate and identify risk factors for infection among children.

## Methods

### Study cohort

We utilized serum samples and survey data collected as part of a longitudinal cohort study in four sites in Kenya from 2014-2018, evaluating the burden of chikungunya virus and dengue virus infections in children [10]. A random subset of 1408 serum samples collected in the last year of the initial cohort study (April 2017 – January 2018) from children 2-18 years of age were included in this analysis. Two geographically distinct areas in Kenya are represented in this analysis: coastal Kenya (Msambweni and Ukunda) and western (Chulaimbo and Kisumu) Kenya. Each geographic region (west and coast) has different baseline infrastructure (higher wealth in the west) and differing weather patterns, with higher temperature and humidity and longer rainy seasons on the coast (Table S1). A rural town was selected in each area (i.e., Msawbweni, Chulaimbo), each with less infrastructure and fewer resources than their adjacent densely populated urban center (i.e., Ukunda, Kisumu) (Table S2) [10]. Households were recruited by random enrollment within confined structured zones in each study community across a similar time period. Serum sample collection and demographic surveys were administered during the same visit. The surveys were designed to collect information about the household, built infrastructure, and behavioral patterns related to mosquito-borne infections, but also captured information relevant to food and waterborne illnesses such as population density and access to piped water and latrines [10]. Serum samples were aliquoted and stored at −70°C until testing. This study protocol was approved by the ethical review boards with the Kenya Medical Research Institute (KEMRI) (SSC95 2611), Stanford University (31488), and Mass General Brigham (2019P000152).

### Serologic Testing

The serum HlyE IgG levels were measured at Mass General by kinetic enzyme-linked immunosorbent assay (ELISA), as previously described [6].

### Statistical Analysis

We estimated seroconversion rates from cross-sectional serosurveys using models of HlyE IgG antibody decay derived from blood-culture confirmed enteric fever cases [6]. These models account for peak antibody responses, decay rates, and variability in immune responses while incorporating multiple biomarkers, measurement noise, and cross-reactivity [11–13]. The approach was implemented using the open-source R package *serocalculator* [14]. The demographic information was paired with the serologic results and analyzed for associations with age, population density, water source, latrine availability, and wealth. The population density was calculated using zonal statistics in software QGIS v3.28.9 (https://qgis.org/en/site/), and population counts were obtained from WorldPop. We divided the population into quartiles representing increasing population density across the cohort. A wealth index was calculated by Multi Correspondence Analysis (MCA), utilizing variables related to tangible assets (e.g., radio, motor vehicle, television, bicycle, telephone), house ownership and characteristics (e.g., number of rooms used for sleeping, people per room, window screens, and building materials), and access to utilities and infrastructure (e.g. source of water, sanitation facility and location) [15,16]. The scores were divided into quartiles representing increasing socioeconomic status (SES) as a measure of wealth throughout the group.[15] All analysis was performed in R (version 4.4.3).

## Results

### Study cohorts

Of the 1408 individuals included in the study, 323 were from Kisumu (west), 323 from Chulaimbo (west), 299 from Ukunda (coast), and 473 from Msambweni (coast) (Table 1). The median age of participants was 10.6 (interquartile range [IQR] 7.8-13.1) years, with the median for each specific community between 10.2-10.9 years of age (Table 1). There was a higher wealth distribution and population density in Kisumu and Ukunda relative to Chulaimbo and Msambweni (Table S1 and Table S2).

**Table 1.** Site characteristics by sex, age, and available variables for piped water source and latrine access.

|  | **Kisumu**<br>(n=323) | **Chulaimbo**<br>(n=323) | **Ukunda**<br>(n=299) | **Msambweni**<br>(n=473) | **Overall**<br>(n=1408) |
| --- | --- | --- | --- | --- | --- |
| Sex – Female | 54% (173) | 46% (146) | 56% (166) | 48% (225) | 50% (719) |
| Age (0-5 years) | 10% (32) | 8% (27) | 7% (20) | 10% (47) | 9% (126) |
| Age (6-10 years) | 44% (140) | 46% (146) | 46% (138) | 48% (227) | 46% (651) |
| Age (11-18 years) | 46% (147) | 46% (145) | 47% (141) | 42% (199) | 45% (632) |
| Median Age (IQR) | 10.2<br>(7.8-12.6) | 10.7<br>(7.8-13.5) | 10.5<br>(7.93-13.1) | 10.9<br>(7.9-13.6) | 10.6<br>(7.8-13.1) |
| Piped Water (1382<br>results available) | >99% (290) | 26% (77) | 92% (259) | 14% (67) | 47% (656) |
| Latrine access<br>(1371 results<br>available) | 100% (301) | 98% (294) | >99% (296) | 41%(193) | 79% (1084) |
| Geographic<br>Location | West | West | Coast | Coast |  |

### HlyE IgG antibody responses

We found higher median anti-HlyE IgG antibody levels in the coastal population (7.73 ELISA units, IQR 4.10-10.97) compared to the western population (0.86 ELISA units, IQR 0.38-2.73), as demonstrated in Figure 1.

**Figure 1.**
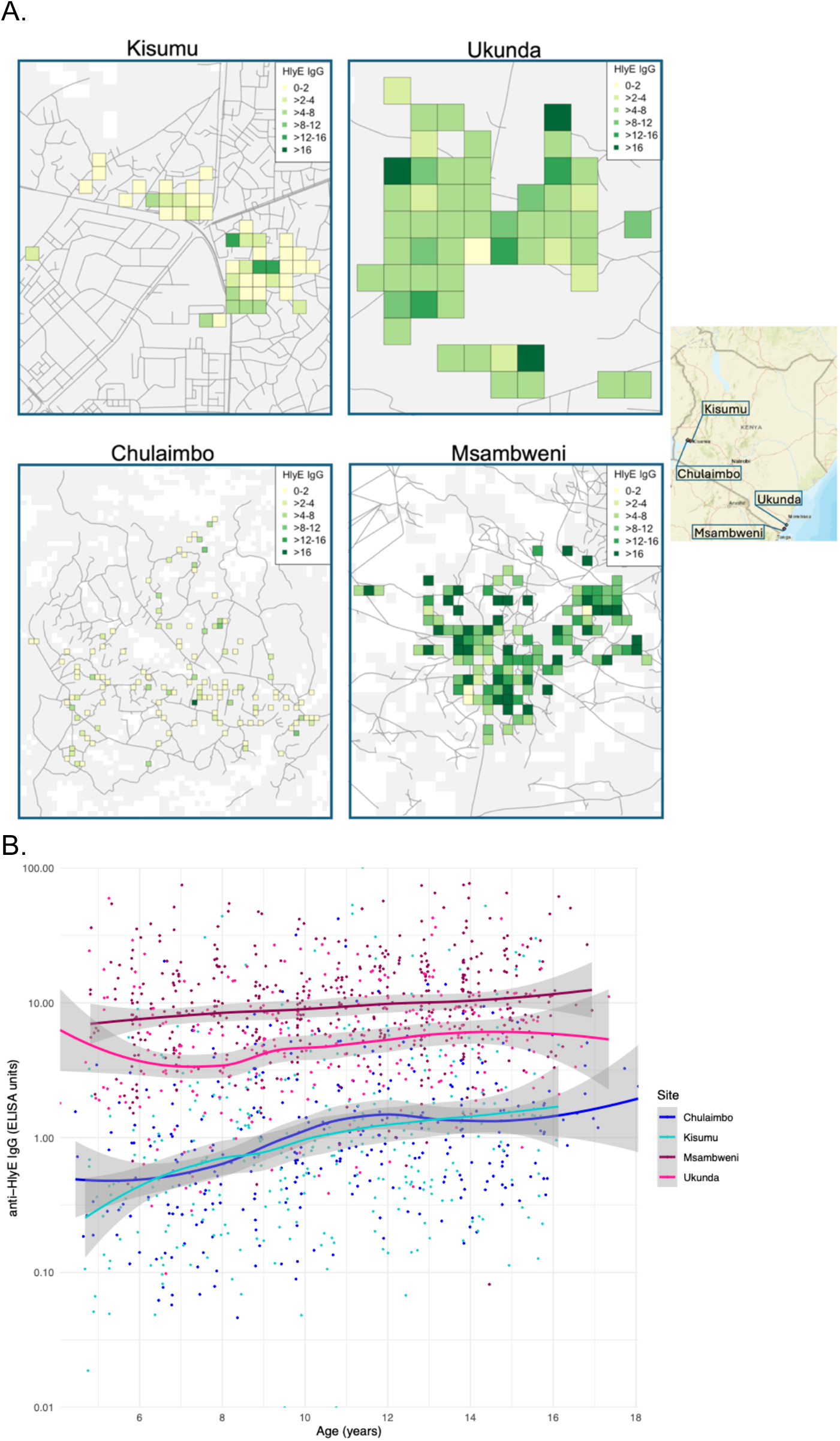
A. Map of Kenya with the respective more densely populated sites, Kisumu and Ukunda, and less densely populated sites, Chulaimbo and Msambweni. The serologic density of anti-HlyE IgG (ELISA units) mapped onto study sites with boxes representing a 100m x 100m grid and colors stratifying by ELISA units. B. Individual antibody responses and smoothed cumulative responses plotted by age by site. Gray shading indicates 95% confidence intervals. A.

### Enteric fever seroincidence rate

The overall seroincidence rate for the Kenyan cohort was 9.1 per 100 person-years (95% CI 8.4-9.8). In the coastal region, the seroincidence rate was 37 per 100 person-years (95% CI 33.8-40.5) and in the western region it was 3.6 per 100 person-years (95% CI 3.0-4.4). There was no significant difference when comparing seroincidence rates by age (less than or greater than 10 years old), however there was a trend of higher seroincidence in the greater than 10 year old group (Figure 2). Piped water, higher wealth, latrine use, and urban location were associated with lower seroincidence rates on the coast (Figure 3). No risk factors were significantly associated with seroincidence rates in the western region of Kenya.

**Figure 2.**
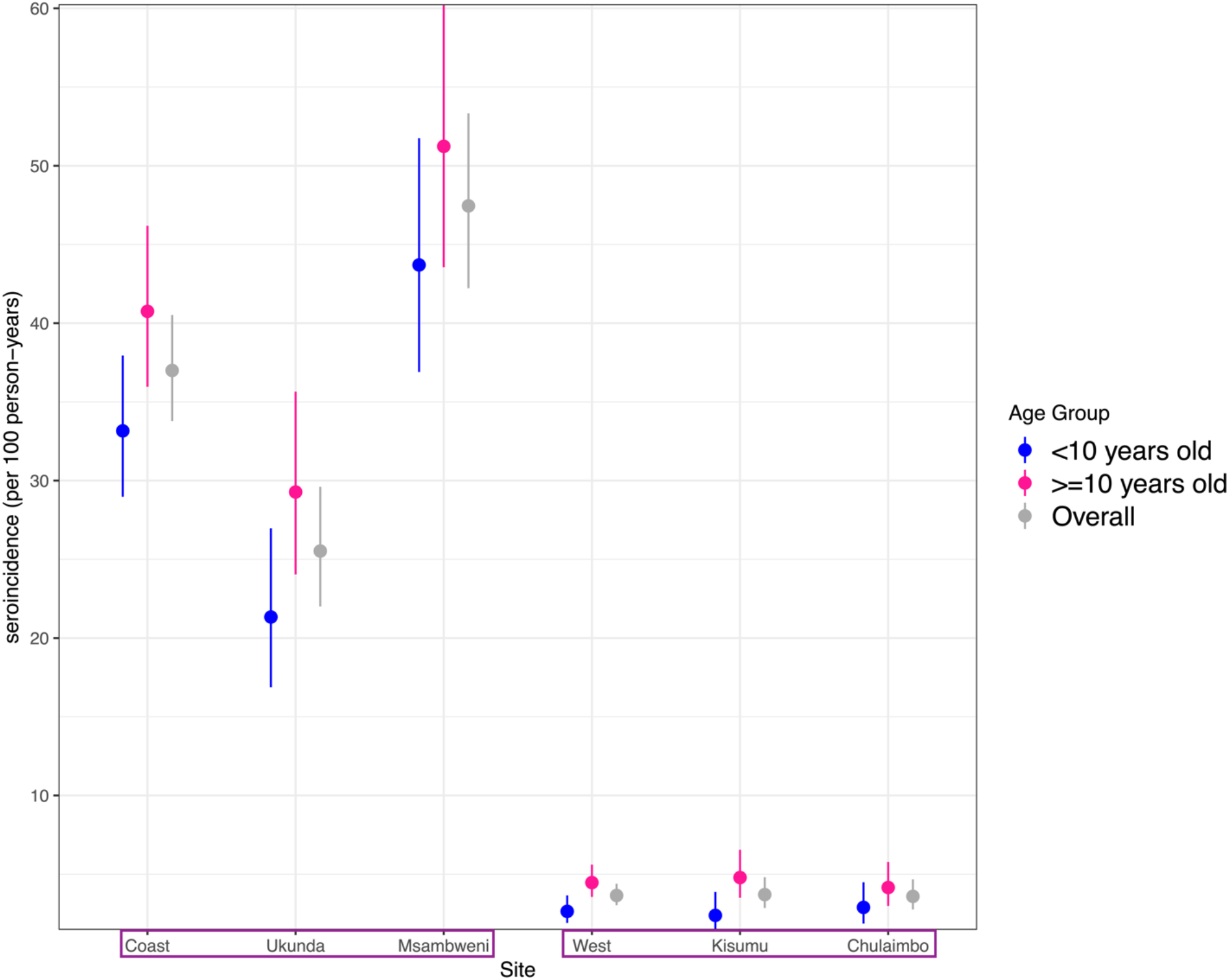
Typhoid seroincidence rate by age (median and 95% confidence interval) and overall distributed by individual site and grouped by west and coast.

**Figure 3.**
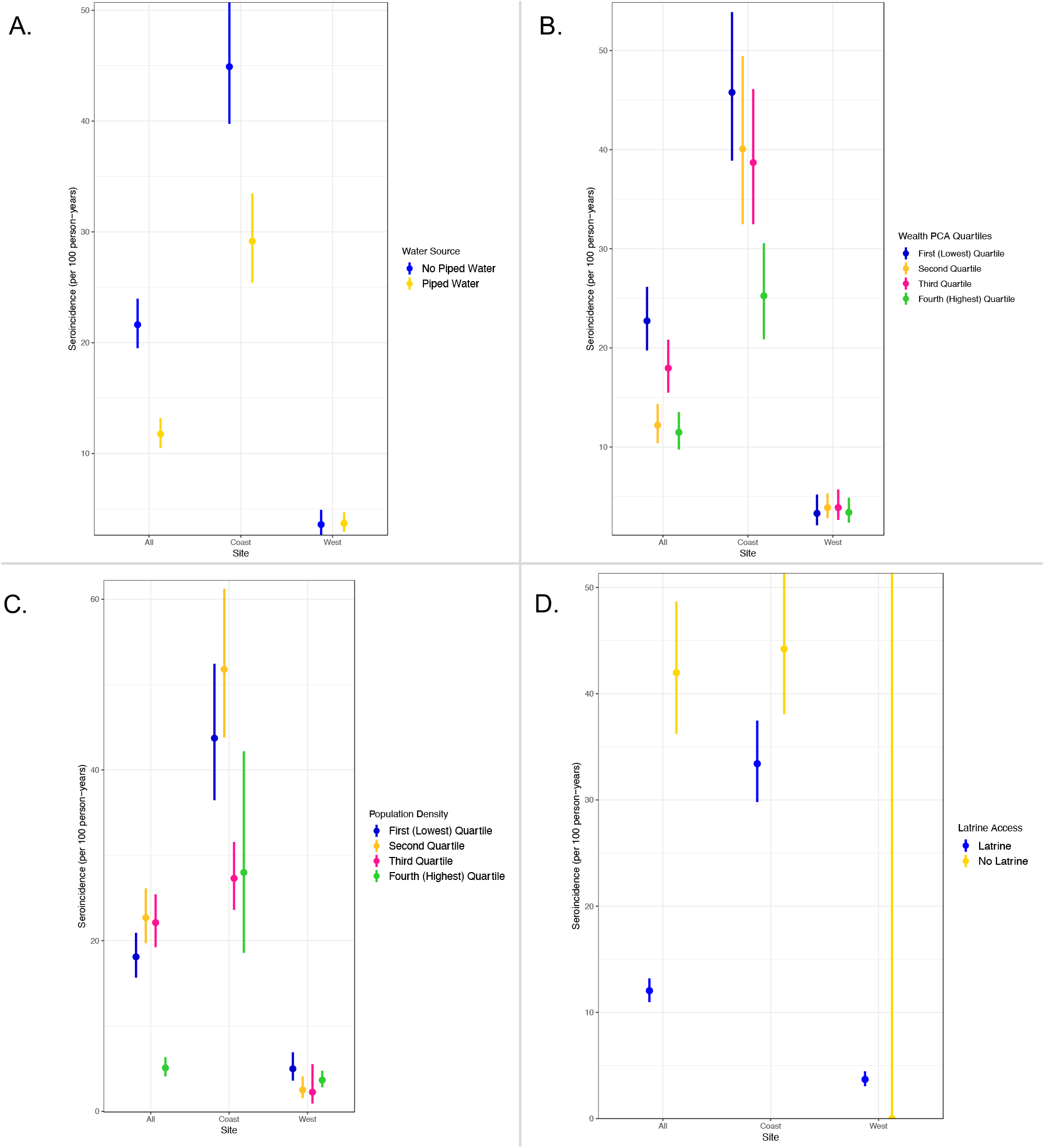
A. Typhoidal Salmonella seroincidence rate by water source. B. Typhoidal Salmonella seroincidence rate by increasing wealth. C. Typhoidal Salmonella seroincidence rate by increasing population density. D. Typhoidal Salmonella seroincidence rate by latrine type.

## Discussion

In this study, we leveraged archived serum samples from a large arbovirus cohort study in Kenya to obtain population-level enteric fever seroincidence rates based on HlyE IgG antibody responses. We found an estimated 10-fold higher seroincidence rate of typhoidal Salmonella on the coast compared to the west. The enteric fever seroincidence rate we estimated in coastal Kenya is similar to the rate estimated in Bangladesh, where ongoing blood culture surveillance for enteric fever has confirmed a higher burden of *Salmonella typhi* and *paratyphi* A [17]. Whereas the seroincidence rate we estimated in Western Kenya is closer to Kathmandu, Nepal (5.8/100 person-years), where the typhoid conjugate vaccine (TCV) campaign was recently launched [6,17].

The enteric fever seroincidence rates estimated here (9,100/100,000 person-years overall) exceed those from previous culture-based studies in the region for which an incidence rate of >100/100,000 cases/year is considered high for typhoid fever. Prior clinical surveillance studies have estimated a range of incidence with an extrapolated crude incidence of 39/100,000 persons per year in Eastern Africa, an adjusted incidence of 284/100,000 person-years of observation in Kibera, Kenya, and an estimated incidence of 620/100,000 person-years in eastern Sub-Saharan Africa [2,3,8,18]. The higher burden estimated by serology results from many factors, including the limited availability and affordability of diagnostics, low sensitivity of culture, and subclinical infections. The variable sensitivity in blood culture sampling and inoculum, especially in young children, renders some febrile individuals falsely negative. Additionally, most typhoidal Salmonella studies evaluate febrile individuals. As patients can be exposed to typhoid without developing a symptomatic infection, it is important to consider that the seroincidence rate will estimate a higher burden than clinical studies by including those with subclinical infection otherwise not captured by clinical surveillance. Furthermore, differences in health-seeking behavior can also account for delayed presentation or missed typhoid cases in various communities [19–21].

In addition to estimating the population-level enteric fever seroincidence rate, we explored potential influence of established risk factors, including population-density, socioeconomic status, and water, sanitation, and hygiene (WASH) measures [22,23]. Consistent with prior studies, we found notable differences in seroincidence rates with water access, latrine use, and overall wealth. On the coast, there was a trend toward a higher burden of enteric fever associated with lower wealth, lower population density, and use of non-piped water. In this study, we found higher enteric fever seroincidence rates in the coastal villages compared to western villages. When evaluating the source of water, the majority of non-piped water was found on the coast, which likely contributed to the higher burden of typhoidal Salmonella found on the coast compared to the western sites. The coastal sites also have higher humidity, relative termperature, and longer rainy seasons, which can be associated with food and waterborne infections [24].

Differences in seroincidence rates were also noted within the coastal sites and in comparison to the western sites. The less densely populated, less developed, and effectively rural site of Msambweni on the coast had the highest seroincidence rate, which deviates from studies in southern Asia and other parts of the world where denser populations have been associated with increased risk of infection. Many of those densely populated communities often do not have access to piped water and live in housing with inadequate sanitation, differing from our study sites [25]. Msambweni had the majority of the lowest SES individuals compared to all four study sites and likely has multiple factors contributing to increased seroincidence rate, including lack of piped water, poor sanitation, and potentially other environmental factors. In contrast, no significant differences were noted in the west between the urban center in Kisumu and its rural adjacent site, Chulaimbo. This was primarily due to lower statistical power to detect a difference given the low seroincidence rate overall in western sites and could also be due to the difference in wealth index, population density, and environmental factors between the geographic sites. There was a greater divide in calculated SES quartiles on the coast compared to the west, where the distribution was closer, as was the calculated seroincidence rate. Lastly, the differing climate and susceptibility to flooding on the coast compared to the west can also contribute to the higher seroincidence rate found on the coast in this study [26,27].

There are several important limitations to this study. The two main risk factors explored in this study (water source and latrine access) were also included in the wealth index calculation and trended in the same direction as wealth, likely influencing the trend identified. Additionally, the original study was focused on mosquito-borne infections and did not include a comprehensive assessment of all the risk factors associated with enteric fever infection. For testing, HlyE IgG captures both *S*. Typhi and *S*. Paratyphi A responses and cannot be differentiated in this study, although *S*. Typhi is presumed to be the dominant pathogen based of blood culture surveillance data.[2] The sample size may not be sufficient to comprehensively represent the greater regions across Kenya. Additionally, although a sample size of 300-400 may be sufficient for calculating the seroincidence rate, these estimations are limited when stratifying by age and other typhoid-associated risk factors. These should be explored further in a larger study. Lastly, these samples were from 2017-2018 and there may be a difference in the seroincidence rate since that time period given climate change, drought/flooding, and other factors like the COVID-19 pandemic where there was a change in movement/behavior.

Despite these limitations, this study demonstrates that the enteric fever seroincidence rate is high in Kenya, particularly on the coast where incidence rates were comparable to other highly endemic areas for typhoid in Asia (e.g. Dhaka, Bangladesh) and over 100 fold higher than estimates by blood culture surveillance. More detailed incidence studies are needed to improve incidence estimates to reveal the comprehensive burden of infection for implementation of public health measures and to determine if the burden remains high in Kenya. These findings suggest that TCV should be considered in coastal and western Kenya in addition to the current practice to provide to high risk groups.

## Data Availability

All data produced in the present study are available upon reasonable request to the authors.

## About the Author

Dr. Khan is a clinical assistant professor in the department of pediatrics and division of infectious diseases at the Stanford University School of Medicine. His research involves studying the epidemiology of mosquito-borne infections and febrile illnesses in children in limited resource settings, with a focus to identify risk factors and mitigating measures in addition to appropriate diagnostics to reduce burden and improve diagnosis of these infections.

## Supplemental Tables

**Table S1.**
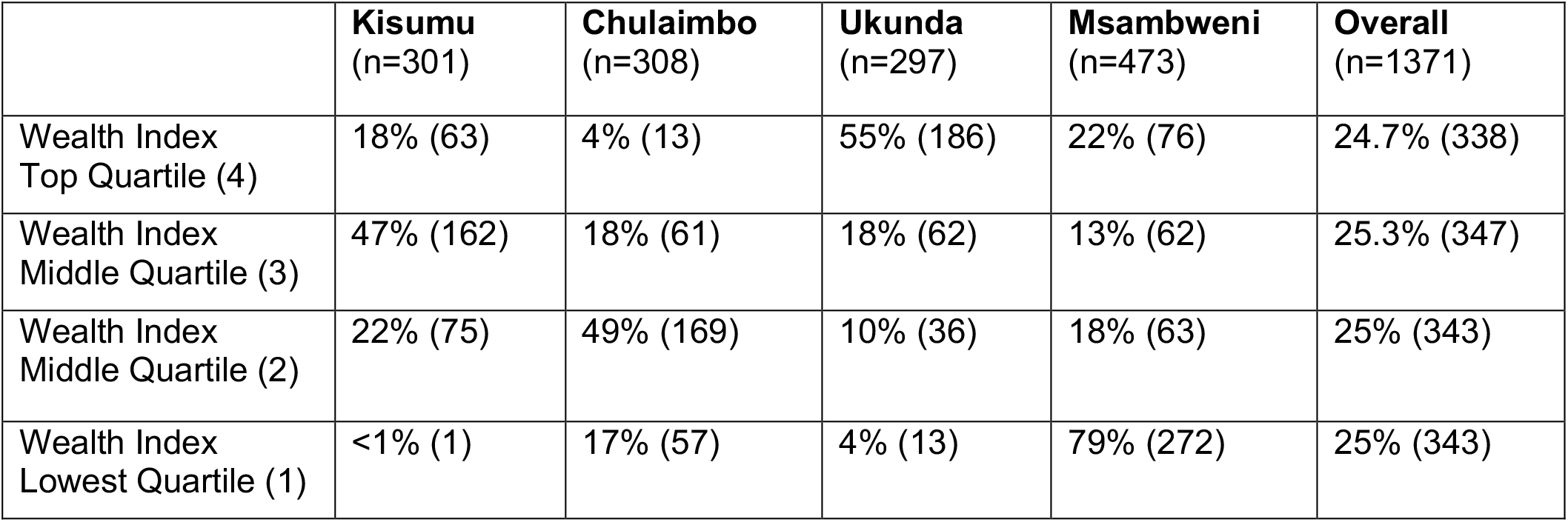
Wealth index with percentages by respective quartile. Factors included in the wealth calculation were house size, house crowding, window screens, bednet ownership, type of floor material, roof type, cookin fuel, water source, light source, land ownership, latrine availability, latrine location, presence of a domestic worker, and ownership of a TV, telephone, radio, bicycle, and motor vehicle.

|  | **Kisumu**<br>(n=301) | **Chulaimbo**<br>(n=308) | **Ukunda**<br>(n=297) | **Msambweni**<br>(n=473) | **Overall**<br>(n=1371) |
| --- | --- | --- | --- | --- | --- |
| Wealth Index<br>Top Quartile (4) | 18% (63) | 4% (13) | 55% (186) | 22% (76) | 24.7% (338) |
| Wealth Index<br>Middle Quartile (3) | 47% (162) | 18% (61) | 18% (62) | 13% (62) | 25.3% (347) |
| Wealth Index<br>Middle Quartile (2) | 22% (75) | 49% (169) | 10% (36) | 18% (63) | 25% (343) |
| Wealth Index<br>Lowest Quartile (1) | <1% (1) | 17% (57) | 4% (13) | 79% (272) | 25% (343) |

**Table S2.** Population density per 100m radius with percentages by respective quartiles.

|  | **Kisumu**<br>(n=303) | **Chulaimbo**<br>(n=308) | **Ukunda**<br>(n=284) | **Msambweni**<br>(n=470) | **Overall**<br>(n=1365) |
| --- | --- | --- | --- | --- | --- |
| Population Density<br>Top Quartile (4) | 90% (301) | (0) | 10% (35) | (0) | 24.6% (336) |
| Population Density<br>Middle Quartile (3) | <1% (2) | 12% (40) | 74% (249) | 14% (47) | 24.8% (338) |
| Population Density<br>Middle Quartile (2) | (0) | 33% (112) | (0) | 67% (232) | 25.2% (344) |
| Population Density<br>Lowest Quartile (1) | (0) | 45% (156) | (0) | 55% (191) | 25.4% (347) |

